# Clinical outcomes of liver failure in patients admitted at the Main Referral Teaching Hospital in Lusaka, Zambia

**DOI:** 10.1101/2025.04.23.25326264

**Authors:** Joseph Phiri, Enock Syabbalo, Amos Hamukale, Paul Kelly, Edford Sinkala

**Affiliations:** Department of Internal Medicine, University of Zambia, Lusaka, Zambia; Department of Internal Medicine, University Teaching Hospital, Lusaka, Zambia; Zambia National Public Health Institute, Lusaka, Zambia

## Abstract

**Background:** Liver failure is a debilitating disease and a major cause of morbidity and mortality. It has a wide spectrum of aetiological factors that lead to different clinical outcomes. Data on the clinical outcomes of liver failure in low income settings are scanty. We set out to evaluate the clinical outcomes and predictors of mortality in liver failure at the main referral and teaching hospital in Zambia.

**Methods:** We consecutively enrolled patients with liver failure at the University Teaching Hospital, Lusaka, Zambia from May 2020 to May 2021. This was an observational cohort study and patients were prospectively followed up for 30 days or until death. Demographics, clinical findings and laboratory investigations were recorded and summary statistics were used to describe data. Predictors of mortality were determined by Cox regression.

**Results:** Out of 59 adult patients whom we evaluated, we enrolled 51 patients who fulfilled the inclusion criteria. The mean age was 43±14 years with 39 (77%) being males. The majority had liver failure whose cause was unknown (27; 53%). Hepatitis B virus was positive in 9 (18%) patients, hepatitis A antibodies were positive in 4 (8%) patients while the antibodies to hepatitis C were positive in 2 (4%) patients. Antituberculosis therapy was suspected to cause liver failure in 9 (18%) patients. Predictors of mortality were low albumin (HR 0.909 [95% CI 0.849-0.973], P=0.006), low neutrophils (HR 0.9928 [95% CI 0.866-0.995], p=0.036), low Karnofsky performance status score (HR 0.417 [95% CI 0.290-0.599], P<0.001), low platelet count (HR 1.002 [95% CI 1.0004-1.00041], P=0.015) and cirrhosis (HR 0.453 [95% CI 0.209-0.979], P=0.044). Mortality rate was 69% (35/51).

**Conclusion:** Patients with liver failure had high mortality rate within the 30-day follow-up. Low albumin, neutrophil count, platelet count and low Karnofsky performance status score including cirrhosis predicted mortality.

## Introduction

Liver failure is one of the major causes of morbidity and mortality worldwide. Acute liver failure (ALF) is life-threatening with mortality rate as high as 50% in the developed countries (1,2). Epidemiological data have indicated that liver failure is a wide, heterogeneous syndrome with associated patterns and outcomes (3,4).

The common causes of liver failure include viral hepatitis, drug toxicity, alcohol abuse and obesity (5). A significant group with indeterminate cause remains despite careful investigations. Lee and others conducted a study in the US and found that the main causes of ALF were drug-induced liver injury (26%), HBV infection (21%) and autoimmune hepatitis (15%) while majority were indeterminate (24%). Prognostic factors were coma and international normalised ratio (INR) at admission. This study found transplant-free survival was improved by N-acetylcysteine only in those with early stage hepatic encephalopathy (5).

Viral hepatitis A, B and E infections remain the common cause of ALF worldwide. While paracetamol-induced hepatotoxicity is the common cause of ALF in the developed countries, anti-tuberculosis therapy is the commonest cause of drug-induced ALF in South Asia and Africa (6). Common causes of chronic liver failure (CLF) are hepatitis B virus (HBV), hepatitis C virus (HCV), alcohol abuse and hepatic steatosis (7). Hepatitis A virus (HAV), HBV and hepatitis E virus (HEV) infections in the setting of chronic liver disease lead to rapid hepatic decompensation and death (7,8). A study conducted in patients with ALF revealed that the independent predictors of mortality were raised bilirubin, prolonged prothrombin time and hepatic encephalopathy grade III and IV (8,9).

Patients who have acute-on-chronic liver disease have poor prognosis. Kumar and others prospectively followed up patients with cirrhosis of varying severity. The mortality rate between HEV infected versus non-infected cirrhosis was 43% versus 22% at 4 weeks, and 70% versus 30% at one year respectively. Patients who acquired acute HBV infection in the setting of chronic liver disease were more likely to suffer decompensation. This was also common in HBV carriers who received immunosuppressive therapy (10).

We have scanty data on the outcome of liver failure in the African setting. We are not sure of the important predictors of mortality in these patients in our region. We therefore set out to evaluate the clinical outcomes and predictors of mortality in liver failure over a period of 30 days at the main referral and University Teaching Hospital (UTH) in Lusaka, Zambia.

## Materials and methods

### Study Design

We conducted an observational prospective study on a cohort of patients with liver failure. We consecutively enrolled patients diagnosed with liver failure over the period of one year from 22nd May 2020 to 21st May 2021.

### Study site and study population

The study was conducted at UTH in the Department of Internal Medicine. The UTH serves as the main referral site for complicated medical and surgical cases in Zambia. The study included adult patients (18 years or older) who were admitted with liver failure in the medical wards. Liver failure was defined as deterioration of liver function evidenced by any two of the following: alanine aminotransferase (ALT) or aspartate aminotransferase (AST) increase of four times or more above the upper limit of normal range (>35iu/L x 4), bilirubin (>50µmol/L), prolonged prothrombin time (or international normalised ratio [INR] ≥1.5), and hepatic encephalopathy (1,2). Subjects were excluded from the study if they had undergone liver surgery or any physical injury to the liver.

### Data Collection and sample size

An interviewer-completed questionnaire was used to obtain clinical and laboratory data of patients with liver failure at the time of inclusion into the study. Written consent was obtained from eligible participants who meet the criteria. However, those who were unable to consent, the next of kin assented for their participation into the study. Vein puncture was done to obtain about 10ml of blood for total bilirubin, ALT, AST, albumin, INR, random blood sugar (RBS), full blood count (FBC), urea, creatinine, human immunodeficiency virus (HIV) and viral hepatitis A, B and C. Basic abdominal ultrasound scan was done. These patients were treated by using the local management protocols. The Karnofsky performance status score (KPS) was done on day 0, 15 and 30 to ascertain the functional status of the patient. Primary outcome was functional improvement measured by KPS while secondary outcome was mortality within follow-up period of 30 days.

### Statistical Analysis

The data collected were analysed by Stata version 13. Categorical variables were summarised as proportions and percentages. Continuous variables were summarised as means and standard deviations when parametric, and median with interquartile range when non-parametric. Paired t-test and Wilcoxon signed-rank test were used to test for differences for continuous variables when parametric or non-parametric, respectively. The chi-squared test was used to test for differences for categorical variable and Fisher’s exact test where the cell was less than 5. Predictors of mortality were established in a multivariate Cox regression. A P-value of less than 0.05 at 95% confidence interval was considered statistically significant.

### Ethical Consideration

The study was approved by the University of Zambia Biomedical Research Ethics Committee (UNZABREC) before the start. Written permission to access the study site and participants was sought from the University Teaching Hospital management.

## Results

A total of 59 patients were considered and assessed for enrolment. Of these, 8 (eight) patients were excluded from the study. Fifty one (51) patients were recruited into the study. There were fewer females (12/51) than males (39/52) who participated in this study with the overall mean age of 43.35±14.10 years. The clinical outcome after 30 days of follow up was mortality of 35 (69%). Eighty per cent of patients were married and more than half had reached secondary level of education. About half had formal employment while approximately one-third were in non-formal employment (Table 4.1).

**Table 4.1:** Baseline demographics of patients with liver failure.

| Variable | Number | Number (%) or Mean (±SD) |
| --- | --- | --- |
| Sex | 51 |  |
| Male |  | 39 (76.50%) |
| Female |  | 12 (23.50%) |
| Age (years) | 51 | 43.35±14.10 |
| Residential area: |  |  |
| Lusaka residents: | 33 | 33 (64.71%) |
| Low density area |  | 3 (5.88%) |
| Medium density area |  | 13 (24.90%) |
| High density area |  | 17 (33.33%) |
| Non-Lusaka residents: | 18 | 18 (35.29%) |
| Marital status: | 51 |  |
| Married |  | 40 (78.43%) |
| Not married |  | 11 (21.57%) |
| Level of education: | 51 |  |
| Primary |  | 4 (7.84%) |
| Secondary |  | 26 (50.98%) |
| Tertiary |  | 21 (41.18%) |
| Occupation: | 51 |  |
| Formal |  | 24 (47.06%) |
| Non-formal |  | 15 (29.41%) |
| None |  | 12 (23.53%) |

The most common clinical feature after jaundice was confusion (98%). The other common clinical features included change in sleep pattern, fever, diarrhoea and itchy skin. Nine patients, 18% (9/51), were on anti-tuberculosis therapy (ATT) at the time of admission. Ten per cent (5/51) reported history of taking herbal medication of which 4 (80%) were non-survivors. Clinical complications included bleeding, hypoglycaemia, seizures, acute renal injury and hepatic encephalopathy. The median functional capacity of these patients determined by KPS was quite low. The median duration of hospital stay was 6 (1–17) days (Table 4.2).

**Table 4.2:** Clinical and radiological characteristics of patients at enrolment.

| Variable | Number | Number (%) or Median (IQR) |
| --- | --- | --- |
| Presenting complaints: | 51 |  |
| Abdominal pain |  | 39 (76.47%) |
| Jaundice |  | 51(100%) |
| Fever |  | 22 (43.14%) |
| Diarrhoea |  | 11 (21.57%) |
| Itchy skin |  | 10 (19.61%) |
| Change in sleep pattern |  | 26 (50.98%) |
| Confused |  | 50 (98.04%) |
| Past medical history: | 51 |  |
| Heart failure |  | 2 (3.92%) |
| Type 2 diabetes mellitus |  | 4 (7.84%) |
| Chronic liver disease |  | 11 (21.57%) |
| HIV |  | 17 (33.33%) |
| Drug history: | 51 |  |
| Paracetamol |  | 9 (17.65%) |
| Anti-tuberculosis therapy (ATT) |  | 9 (17.65%) |
| Antiretroviral therapy (cART) |  | 17 (33.33%) |
| Herbal medication |  | 5 (10.00%) |
| Alcohol |  | 29 (56.86%) |
| Physical/radiological examination: | 51 |  |
| Mean arterial pressure (MAP) |  | 89.86±14.01 |
| Pulse (beats/min) |  | 101.35±14.24 |
| Axillary temperature (°C) |  | 37.2 IQR,(36.50-37.80) |
| BMI |  | 24.08 IQR (21.08-26.53) |
| Pallor |  | 21 (41.18%) |
| Abdominal tenderness |  | 27 (52.94%) |
| Hepatomegaly |  | 25 (49.02%) |
| Fatty liver |  | 6 (11.76%) |
| Cirrhosis |  | 18 (35.29%) |
| Ascites |  | 28 (54.90%) |
| Splenomegaly |  | 6 (11.76%) |
| Pedal oedema |  | 21 (41.18%) |
| Acute liver failure (ALF) | 51 | 12 (23.53%) |
| Acute-on-chronic liver failure/ chronic liver failure (ACLF/CLF) | 51 | 39 (76.47%) |
| Clinical complications at admission: | 51 |  |
| Bleeding | 12 | 12 (23.53%) |
| Hypoglycaemia (<3.00mmol/L) | 7 | 7 (13.73%) |
| Seizures | 4 | 4 (7.84%) |
| Renal failure | 26 | 26 (50.98%) |
| Hepatic encephalopathy: | 51 |  |
| Grade I-II |  | 30 (58.82%) |
| Grade III-IV |  | 21 (41.18%) |
| Karnofsky performance status score (%) | 51 | 30 IQR (10-50) |
| Hospital stay (days) | 51 | 6 IQR (1-17) |

Leucocytosis, thrombocytopaenia and anaemia were common in our patients. The median total bilirubin, international normalised ratio (INR) and transaminases were high while the mean serum albumin was low. The cause of liver failure (53%) was unknown in majority of patients due to inadequate resources. About a third (18%; 9/51) had HBV and ATT (anti-tuberculosis therapy) respectively as the cause of liver failure followed by HAV (8%; 4/51) while about 4% (2/51) was due to probable HCV. One (2%) patient had HAV/HBV co-infection while the other patient had HBV/HCV co-infection. One-third of patients were infected by HIV. Four per cent of the patients had HIV/HAV, 4% (2/51) had pulmonary tuberculosis/HAV, 2% (1/51) had HIV/HBV and 2% (1/51) had HIV/HCV co-infections (Table 4.3).

**Table 4.3:** Baseline laboratory characteristics at admission.

| Variable | Number | Number (%) or Median (IQR) |
| --- | --- | --- |
| Laboratory tests: | 51 |  |
| White cell count (x10<sup>9</sup>/L) |  | 9.64 (2.14-22.94) |
| Haemoglobin (g/dL) |  | 10.91±2.93 |
| Mean corpuscular volume |  | 85.50 (77.6-92.20) |
| Platelets (x10<sup>9</sup>/L) |  | 141 (28-404) |
| Neutrophils |  | 7.07 (1.21-18.3) |
| Sodium (mmol/L) |  | 135.12±8.03 |
| Potassium (mmol/L) |  | 3.97 (3.02-4.90) |
| Urea (mmol/L) |  | 7.09 (3.33-18.10) |
| Creatinine (μmol/L) |  | 106.10 (27.7-606) |
| Random blood sugar (mmol/L) |  | 5.6 (4.0-8.0) |
| Total bilirubin |  | 242.30 (58-513.2) |
| INR |  | 2.10 (0.92-4.5) |
| ALT (IU/L) |  | 267 (53-1243) |
| AST (IU/L) |  | 422.1 (45-1938) |
| Albumin (g/L) |  | 23.46±6.35 |
| HAV | 51 | 4 (7.84%) |
| HBV | 51 | 9 (17.65%) |
| Probable HCV | 51 | 2 (3.92%) |
| HIV | 51 | 17 (33.33%) |

All variables are expressed as median (IQR) unless stated otherwise (mean±SD). HAV – Hepatitis A virus, HBV – Hepatitis B virus; HCV – Hepatitis C virus, HIV – Human immunodeficiency virus; INR – International normalised ratio; ALT – Alanine aminotransferase; AST – Aspartate aminotransferase

Predictors of mortality by Cox regression analysis were low serum albumin, neutrophil count, platelet count and KPS, and cirrhosis (Table 4.4).

**Table 4.4:** Multivariate analysis by Cox regression.

|  | **Hazard ratio** | **Standard error** | **P value** | **95% Confidence interval** |
| --- | --- | --- | --- | --- |
| Albumin | 0.9089964 | 0.0317008 | 0.006 | 0.8489398-0.9733015 |
| Neutrophils | 0.9282716 | 0.0330087 | 0.036 | 0.8657787-0.9952753 |
| KPS score | 0.4166237 | 0.0771699 | 0.000 | 0.2897868-0.5989759 |
| Platelets | 1.002263 | 0.0009293 | 0.015 | 01.000444-1.0004086 |
| Cirrhosis | 0.4526047 | 0.1781577 | 0.044 | 0.2092486-0.9789836 |

KPS – Karnofsky Performance Status score

The proportion of patients with advanced hepatic encephalopathy, seizures, hypoglycaemia, high serum total bilirubin, ALT, INR and low albumin was higher among non-survivors than survivors. Eighty per cent of the patients who had taken herbal medications and ATT were non-survivors. Most of the patients who died had ALF followed by those who had acute-on-chronic liver failure (Table 4.5).

**Table 4.5:** Baseline variables of survivors and non-survivors.

| Variable | Survivors (n=16) | Non-survivors (n=35) | P value |
| --- | --- | --- | --- |
| Sex, women: men | 5:11 | 7:28 | 0.039 |
| Age (years) | 44.04±16.45 | 43.03±13.14 | 0.810 |
| Participants: |  |  |  |
| Lusaka residents | 13 (81.25%) | 20 (57.14%) | 0.031 |
| Non-Lusaka residents | 3 (18.75%) | 15 (42.86%) | 0.230 |
| Alcohol use, n (%) | 9 (56.25%) | 20 (57.14%) | 0.103 |
| Anti-retroviral therapy (cART) | 2 (12.50%) | 15 (42.86%) | 0.033 |
| Clinical features: |  |  |  |
| Abdominal tenderness | 10 (62.50%) | 17 (48.57%) | 0.360 |
| Hepatomegaly | 7 (43.75%) | 19 (54.29%) | 0.494 |
| Ascites | 8 (50.00%) | 28 (80.00%) | 0.029 |
| Cirrhosis | 5 (31.25%) | 13 (37.14%) | 0.690 |
| Karnofsky performance score | 40±10% | 30±10% | 0.001 |
| Hospital stay | 6.76±2.00 | 6.20±5.96 | 0.250 |
| Acute liver failure, n (%) | 4 (25.00%) | 8 (22.86%) | 0.620 |
| ACLF/ CLF, n (%) | 12 (75.00%) | 27 (77.14%) | 0.049 |
| Hepatic encephalopathy, n (%): |  |  |  |
| Grade I-II | 14 (87.50%) | 16 (45.71%) | 0.004 |
| Grade III-IV | 2 (12.50%) | 19 (54.29%) | 0.004 |
| Complications (%): |  |  |  |
| Bleeding | 4 (25.00%) | 7 (20.00%) | 0.870 |
| Renal failure | 7(43.75%) | 19 (54.29%) | 0.232 |
| Laboratory investigations: |  |  |  |
| Neutrophils (x10<sup>9</sup>/L) | 7.40(5.82-14.45) | 6.36(3.51-11.35) | 0.302 |
| Haemoglobin (g/dL) | 10.93±3.09 | 10.89±2.90 | 0.970 |
| Platelet (x10<sup>9</sup>/L) | 126.5(47-173) | 141(104-225) | 0.068 |
| INR | 1.95(1.11-2.55) | 2.20(1.90-3.60) | 0.049 |
| Total bilirubin (µmol/L) | 165.6(95.7-295.7) | 252.6(185.2-424.8) | 0.027 |
| ALT (IU/L) | 220.3(190.9-441.4) | 275.1(201-625) | 0.500 |
| AST (IU/L) | 361.3(290.7-724.1) | 478(295-973.8) | 0.560 |
| Albumin (g/L) | 27.86±6.43 | 21.46±5.28 | 0.000 |
| HBsAg – positive | 2 (12.50%) | 7 (20.00%) | 0.521 |

**Key:** All variables are expressed as median (IQR) unless stated otherwise (mean±SD). ACLF – Acute-on-chronic liver failure; CLF – Chronic liver failure ALT – alanine aminotransferase; AST – Aspartate aminotransferase; INR – International normalised ratio; HBsAg – Hepatitis B surface antigen

## Discussion

This study identified some important early predictors of mortality in patients with liver failure in an African setting. The multivariate analysis showed that low serum albumin, neutrophils, Karnofsky Performance Status scale (KPS), and platelets, and cirrhosis were independent predictors of mortality. This calls for prompt evaluation of these blood markers in patients with liver failure in order to consider replacement therapy where necessary. Fernadendes and others also noted that hypoalbuminemia predicts mortality in patients with liver failure especially those with cirrhosis (11). In decompensated cirrhosis most of the synthetic functions of the liver are impaired. Studies advocating for the use of albumin infusion in patients with cirrhosis have conflicting results. A trial of albumin in patients with spontaneous bacterial peritonitis showed some benefit (12). A study by Louise and colleagues showed no benefit of albumin infusion in patients with cirrhosis in the United Kingdom (13). Data in an African setting like Zambia are scanty on use of albumin in liver failure. Therefore, this study lays some ground to advocate for clinical trials of albumin use in liver failure.

In our study a quarter of non-survivors had history of bleeding which could have been due to thrombocytopenia and coagulopathy. Thrombocytopaenia in cirrhosis is usually as a result of hypersplenism and reduced production of thrombopoietin, a hormone important for production of platelets (7,14). Cirrhosis being the end stage liver disease needs evaluation for possible consideration of liver transplantation in good time.

Leucocytosis and development of neutropenia or neutrophilia are predictors of mortality in liver failure. Vaquero and colleagues showed patients with liver failure are prone to bacterial infection as a result of depressed immunity due to decreased synthesis of immune factors (15). Our study had patients with cirrhosis which can complicate spontaneous bacterial peritonitis (SBP). This could have led to abnormal neutrophil count. The lack of immune factors in these patients with sepsis could suggest that undiagnosed infection might be the cause of abnormal neutrophil levels. This could explain poor prognosis in our study where more than two-thirds of the patients with low neutrophil levels died.

Ascites is one of the most common complications of cirrhosis along with hepatic encephalopathy, hepatorenal syndrome and upper gastrointestinal bleeding (13). In our study, more than half of the patients presented with ascites. This was similar to previous studies which found about 60% of the patients with cirrhosis would develop ascites within 10 years after diagnosis (13,16). Planas and others found that patients with cirrhosis who had ascites were at a high risk of developing various complications of liver disease, including SBP and hepatorenal syndrome (17).

In our study, all patients presented with evidence of liver parenchymal injury with elevated ALT and AST. The high mortality rate despite the provision of treatment could be associated with late presentation to our hospital.

In our study, the prevalence of HBV was higher (18%) as compared to the national prevalence of 5.6% (18). The higher prevalence of HBV is likely due to bias since this was a hospital based study. The patients with HBV had a high mortality rate of 80% which could be attributed to lack of initiating treatment for HBV in good time.

Evaluating drug history in patients with liver failure is important. Our study found that anti-tuberculosis therapy (ATT) was an important cause of liver injury in 18% of the patients with liver failure admitted to our hospital. The underlying cause of liver injury in these patients maybe idiosyncratic reaction to ATT rather than dose-related toxicity (8,19).

Our study also showed that employing a monitoring tool in the follow-up of patient with liver failure could be of value. Karnofsky Performance Status score (KPS) of 30 was associated with increased mortality of more than 50% in our patients. Tandon and colleagues also found that low KPS was associated with hepatic encephalopathy, longer hospital stay, leucocytosis and higher MELD score at discharge (20). A study done by Khalid and others in Pakistan showed that KPS can be utilized to identify patients with cirrhosis who are at risk of three-month post discharge mortality (21).

Our study was met with some limitations. Due to lack of availability of liver transplantation facility in our hospital, a decision for early enrolment for the life-saving treatment option could not be offered to these patients. This could have helped to identify predictive parameters for the disease progression and mortality better. Considering all-cause mortality was used as one of the end-points and that the study protocol did not include post-mortem, it is possible that some deaths may not have been caused by liver failure. A larger sample size could help in better identifying the predictors of mortality. Therefore, large prospective studies could be worthwhile in addressing these challenges.

## Conclusion

Our patients with liver failure had a high mortality rate within the 30 day follow-up. Low albumin, neutrophil count, platelet count and low Karnofsky performance status score including cirrhosis predicted mortality.

## Data Availability

All relevant data are within the manuscript and its supporting information files

## Acknowledgements

We wish to acknowledge the nurses, interns, registrars, senior registrars and consultants at the UTH who helped in the management of the cases recruited for this study. We also thank the Department of Internal Medicine and management of the hospital for giving us access to these cases.

## Notes

### Competing Interest Statement

The authors have declared no competing interest.

### Funding Statement

The author(s) received no specific funding for this work.

### Author Declarations

IRB: Universit of Zambia Biomedical Research Ethics Committee (UNZABREC) Approval number: REF: 660-2019 The approval was base on the following documents that were submitted for review a) Study proposal b) Questionnaires c) Participant Consent Form Form of consent: Written Date of approval: May 22nd, 2020

